# Genetic Risk for Hemochromatosis is Associated with Movement Disorders

**DOI:** 10.1101/2021.08.16.21262117

**Authors:** Robert Loughnan, Jonathan Ahern, Cherisse Thompkins, Clare E. Palmer, Leo Sugrue, John Iversen, Wesley Thompson, Ole Andreassen, Terry Jernigan, Anders Dale, Mary ET Boyle, Chun Chieh Fan

## Abstract

Hereditary hemochromatosis (HH) is an autosomal recessive genetic disorder that can lead to iron overload, causing oxidative damage to affected organs. HH type 1 is predominantly associated with homozygosity for the mutation p.C282Y. Previous case studies have reported tentative links between HH and movement disorders, e.g., Parkinson’s disease, and basal ganglia abnormalities on magnetic resonance imaging. We investigated the impact of p.C282Y homozygosity: on whole brain T2 intensity differences, a measure of iron deposition, and; on measures of movement abnormalities and disorders within UK Biobank. The neuroimaging analysis (154 p.C282Y homozygotes, 595 matched controls) showed that p.C282Y homozygosity was associated with decreased T2 signal intensity in motor circuits (basal ganglia, thalamus, red nucleus, and cerebellum; Cohen’s d > 1) consistent with substantial iron deposition. Across the whole UK Biobank (2,889 p.C282Y homozygotes, 496,968 controls), we found a significant enrichment for movement abnormalities in male homozygotes (OR (95% CI) = 1.82 (1.27-2.61), p=0.001), but not females (OR (95% CI) = 1.10 (0.69-1.78), p=0.71). Among the 31 p.C282Y homozygote males with a movement disorder only 7 had a concurrent HH diagnosis. These findings indicate susceptibility to iron overload in subcortical structures in p.C282Y homozygotes, and confirmed an increased risk of movement abnormalities and disorders in males. Given the effectiveness of early treatment in HH, screening for p.C282Y homozygosity in high risk individuals may offer a potential avenue to reduce iron accumulation in the brain and limit additional risk for the development of movement disorders among males.

## Introduction

Hereditary hemochromatosis (HH) is a disorder that leads to iron overload in the body. HH type 1 is predominantly related to a HFE gene mutation, with 95% of cases being homozygote for p.C282Y (p.Cyst282Tyr) mutation^1^. The excess iron absorbed by the body leads to an accumulation of iron in organs, particularly in the liver leading to increased risk for liver disease and diabetes^2^. With a homozygosity rate of approximately 0.6% in northern European populations, it has been deemed the most prevalent genetic disorder in Europe^3,4^. The penetrance of HH and other associated diseases in p.C282Y homozygote individuals appears to be larger for males than females^5^, leading researchers to believe that expelling excess iron through menstruation and pregnancy lowers disease burden and penetrance in females. The primary treatment for HH is phlebotomy which appears to be effective at reducing adverse clinical outcomes^3^ if started early. As a result, some researchers have advocated for re-evaluating screening and early case ascertainment^2^.

Although the impact of HH and p.C282Y homozygosity on the liver and heart is largely accepted^2,3^, its effect on the central nervous system is still disputed. While some studies have reported a higher risk for Alzheimer’s and Parkinson’s disease in p.C282Y homozygote individuals^5,6^, other studies have reported no risk^7,8^ or a protective effect^9^. These conflicting results may be explained by modest sample sizes and pooled analysis across sexes. Case studies of HH individuals experiencing neurological deficits have confirmed neuroimaging abnormalities in the basal ganglia, substantia nigra, and cerebellum^10,11,12^, all regions known to have a substantial role in the control of movement^13^. However, these previous studies included only individuals diagnosed with HH, and therefore do not describe neurological deficits and abnormalities of p.C282Y homozygote individuals independent of HH diagnosis. A recent study performed an analysis of brain MRI data from 206 p.C282Y homozygote individuals taken from the UK Biobank^14^. Using analyses of T2^*^ signal in predefined anatomical regions (Region Of Interest-based) they found indications of increased iron deposition in subcortical structures and cerebellum for p.C282Y homozygote individuals, and increased risk of dementia in p.C282Y homozygote males.

Since the implicated brain regions with p.C282Y homozygosity are known to have a strong involvement in motor functions^15^ and reports of movement deficits in HH individuals^5,6,10,11^, we investigated the relationship between p.C282Y homozygosity and movement disorders in 502,536 individuals from the UK Biobank, with improved imaging technology enabling greater granularity of associations across the brain. Specifically, we investigated: i) the impact of p.C282Y homozygosity on whole brain voxel-wise measures of iron deposits; ii) the sex stratified association of p.C282Y homozygosity with movement disorders and tested for overlap with HH diagnosis.

## Methods

### UK Biobank Sample

Genotypes, MRI scans, demographic and clinical data were obtained from the UK Biobank under accession number 27412, excluding participants who withdrew their consent. This resulted in a total sample of 502,536 individuals with a mean age of 57.0 years (standard deviation 8.1 years), 229,134 male. We used UK Biobank v3 imputed genotype data^30^. From this sample, 2,889 individuals were identified as homozygote for p.C282Y (A/A at rs1800562), 1,293 male. As previous research does not indicate intermediate disease burden for p.C282Y heterozygotes^2^, we coded control individuals as homozygote for no risk allele (G/G at rs1800562) or heterozygote (A/G at rs1800562) i.e., with a recessive model of inheritance. This resulted in 499,647 controls, 227,841 male.

### Neuroimaging Analysis

#### Image acquisition

T1 weighted and diffusion weighted scans were collected from three scanning sites throughout the United Kingdom, all on identically configured Siemens Skyra 3T scanners, with 32-channel receiver head coils. For diffusion scans, multiple scans with no diffusion gradient were collected (b=0 s/mm^2^) to fit diffusion models. The average of these b=0 scans was used as voxel-wise measures of T2 intensities. Diffusion weighted scans were collected using an SE-EPI sequence at 2mm isotropic resolution. T1 scans were collected using a 3D MPRAGE sequence at 1mm isotropic resolution.

#### Image Preprocessing

Scans were corrected for nonlinear transformations provided by MRI scanner manufacturers^31,32^, and T2 images were registered to T1 weighted images using mutual information^33^. Intensity inhomogeneity correction was performed by applying smoothly varying, estimated B1-bias field^34^. Images were rigidly registered and resampled into alignment with a pre-existing, in-house, averaged, reference brain with 1.0 mm isotropic resolution^34^.

#### Atlas Registration

To allow for voxel-wise analysis, subjects’ imaging data were aligned using a multimodal nonlinear elastic registration algorithm. At the end of the preprocessing steps outlined in *Image Processing* and described in detail in Hagler et al. ^34^, subjects’ structural images and diffusion parameter maps were aligned to an UK Biobank-specific atlas, using a custom diffeomorphic registration method.

#### Labelling regions of interest (ROI)

Subcortical structures were labeled using Freesurfer 5.3^35^. Subjects’ native space Freesurfer parcellations were warped to the atlas space and averaged across subjects. Additional subcortical nuclei, not available in the FreeSurfer segmentation, were labeled by registering readily available, downloadable, high spatial resolution atlases to our atlas space. The Pauli atlas was generated using T1 and T2 scans from 168 typical adults from the Human Connectome Project (HCP)^36^. The Najdenovska thalamic nuclei atlas was generated using a k-means algorithm taking as inputs mean fibre orientation density spherical harmonic coefficients from within a Freesurfer parcellation of the thalamus, using adult HCP data from 70 subjects^37^. All subcortical ROIs and abbreviations are listed in Supplementary Table 2.

#### Covariate Matched Controls

From the full 2,889 p.C282Y homozygotes, only 154 had qualified imaging. As we did not want to have a large imbalance between the number of controls and p.C282Y homozygotes, we selected covariate matched controls at a ratio of 4:1 (controls:cases). We matched controls on age, sex, scanner, mean cortical area, and top ten components of genetics ancestry. These matched controls were generated through using the sample of 154 individuals with qualified imaging, fitting a logistic model to predict p.C282Y homozygosity from the listed covariates, and then generating propensity scores^38^ for each individual. Controls were selected on having propensity scores with a threshold of |*S*_1_ − *S*_2_ | ≤ *threshold*, where *S*_1_ and *S*_2_ were the respective propensity scores of a p.C282Y homozygote and their matched control. The threshold was set to 1.5×10^−4^. We performed this procedure by using the *pymatch* library (version 0.3.1) in python^39^. This resulted in 154 p.C282Y homozygotes (64 male) and 595 controls (248 male).

#### Statistical analysis

General linear effect models were applied univariately to test the association between p.C282Y homozygosity and T2 intensities. Each voxel-wise T2 intensity was pre-residualized for age, sex, scanner, and top ten principal components of genetic ancestry. We then calculated Cohen’s d effect sizes as the residualized voxel-wise differences between p.C282Y homozygotes and controls. As previous research has indicated a higher disease burden for p.C282Y homozygote males vs females, we in supplementary results we additionally performed a sex-stratified analysis in 312 males (64 p.C282Y homozygote) and 437 females (90 p.C282Y homozygote).

### Neurological Disease Burden Analysis

Given our neuroimaging findings of substantially lower T2 intensities of p.C282Y homozygotes in motor and gait circuits of the brain – indicative of iron deposition – we wanted to test if p.C282Y homozygosity imparted any risk for i) movement disorders, ii) gait disorders and iii) a broad category of neurological disorders. As completing imaging was not an inclusion criterion for this portion of the analysis, we included the entire sample listed above. We did not perform any covariate-matching of controls. Sex stratified logistic models fit 229,134 males (1,293 p.C282Y homozygote) and 273,402 females (p.C282Y 1,596 homozygote) to predict diagnosis from p.C282Y homozygosity status controlling for age, sex, and top ten principal components of genetic ancestry. We fit three models for each sex to test the domains described above, predicting: movement disorders [ICD10: G20-G26], abnormalities of gait and mobility [ICD10: R26], and other disorders of the nervous system [ICD10: G90-99]. We fit an additional three models, two to for the largest diagnoses of movement disorders; Parkinson’s disease [ICD10: G20] and essential tremor [ICD10: G25], as well as a super-category of significantly associated diagnoses combining a diagnosis of either a) movement disorders or b) other disorders of the nervous system into a single outcome.

## Results

### Neuroimaging Analysis

We performed a voxel-wise analysis of T2 intensities (lower T2 intensity consistent with higher iron deposition^17^) of using p.C282Y homozygosity status as our predictor of interest – see Table 1 for sample sizes. We found evidence for iron deposition reflected in substantially lower T2 intensities localized to the basal ganglia, thalamus and cerebellum in p.C282Y homozygote individuals. p.C282Y homozygosity was associated with lower T2 intensities in bilateral caudate nucleus, putamen, thalamus (specifically the ventral anterior, ventral lateral dorsal, ventral-lateral ventral and pulvinar nuclei), red nucleus, sub-thalamic nucleus, and both white and grey matter of the cerebellum (Figure 1). Additionally, we observed higher T2 intensities in the white matter of the superior cerebellar peduncle, possibly indicating gliosis in this region^18^ which comprises the primary output pathway from the cerebellum to the thalamus and red nucleus. Supplementary sex-stratified analysis revealed similar effects in males and females, with p.C282Y homozygote females having on average approximately 30% smaller effects than males (supplementary figures 1-2).

**Table 1.** Sample size for each analysis.

|  | *Neuroimaging Analysis* | *Neurological Disease Analysis* |
| --- | --- | --- |
| *Sample Size* | 749 (312 males) | 502,563 (229,134 males) |
| *p.C282Y Homozygotes* | 154 (64 males) | 2,889 (1,293 males) |

**Figure 1.**
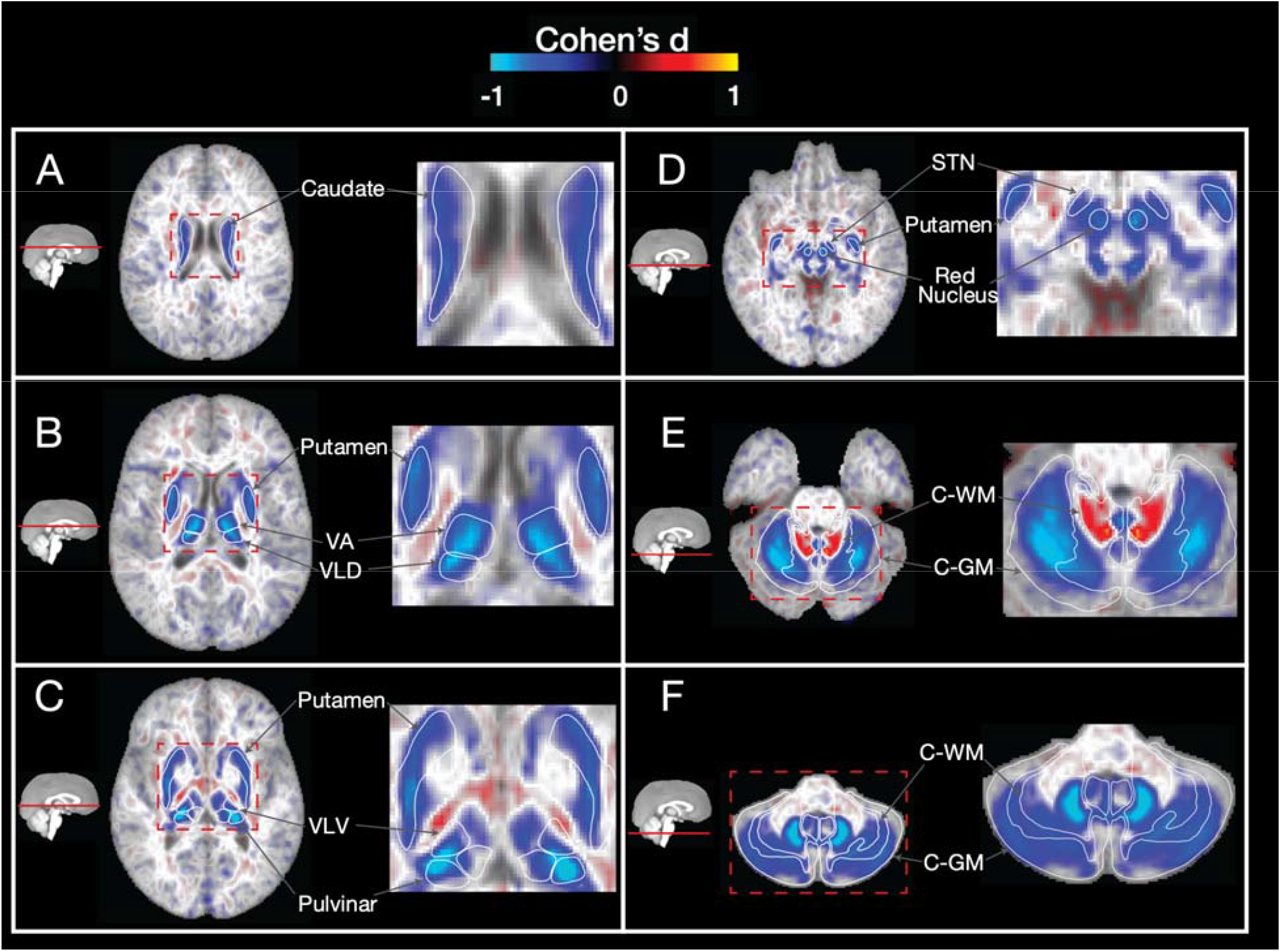
Voxelwise associations (cohen’s d) of T2 intensities between matched controls vs p.C282Y homozygote individuals. Blue regions represent lower T2 intensities (indicating higher iron deposition) for p.C282Y homozygotes. Lower T2 intensities are observed for p.C282Y homozygotes in the A. caudate nucleus, B. putamen, ventral anterior and ventral lateral dorsal nuclei of the thalamus, C. ventral-lateral ventral and pulvinar nuclei of the thalamus, D. red nucleus, sub thalamic nucleus, E. and F. grey and white matter of the cerebellum. Higher T2 intensities are observed in E. in the superior cerebellar peduncle (primary output pathway connecting the cerebellum to the thalamus and red nucleus).

### Neurological Disease Burden Analysis

The brain regions identified in our analysis have previously been implicated to have a strong involvement in motor control^15,19^. We aimed to determine if p.C282Y homozygote individuals had an enrichment for movement abnormalities, including movement diseases and gait/mobility, or other disorders of the nervous system. Table 1 displays the sample size for this analysis. As males appear to have a greater penetrance for HH and other associated diseases^2^, we performed a sex-stratified analysis. We found that p.C282Y homozygote males had a higher chance of being diagnosed with a movement disorder (OR (95% CI) = 1.82 (1.27-2.61), p=0.001) and other disorders of the nervous system (OR (95% CI) = 1.51 (1.06-2.14), p=0.02). The International Classifications for Diseases (ICD) chapter of movement disorders includes Parkinson’s disease and essential tremor, which were both associated with p.C282Y homozygosity in males (Parkinson’s disease: OR (95% CI) = 1.78 (1.14-2.79), p=0.01 and essential tremor: OR (95% CI) = 1.92 (1.03-3.60), p=0.04). p.C282Y homozygote males did not have a higher chance of being diagnosed with gait or mobility disorders (OR (95% CI) = 0.96 (0.65-1.42), p=0.85). No significant associations were found for p.C282Y homozygote females for any diagnosis tested – see Figure 2 panel A and Supplementary Table 2.

**Figure 2.**
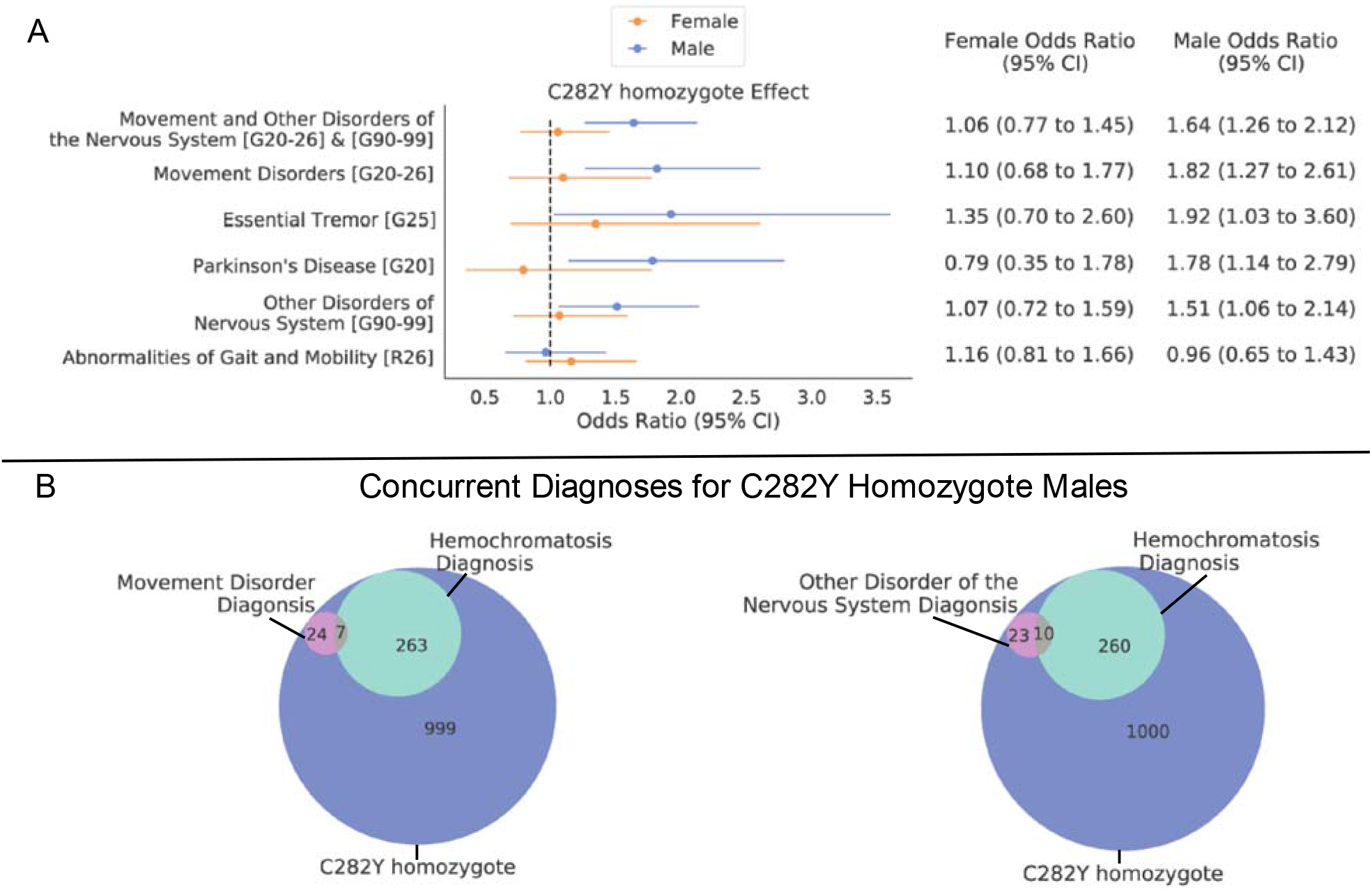
A. sex-stratified effect of C282Y homozygosity for neurological disorders (y axis), values in square brackets indicate ICD10 codes. Dotted vertical line indicates an odds ratio of 1 i.e., null effect. B. Venn diagrams indicating diagnosis overlap for C282Y homozygote males of hemochromatosis and i) movement disorders or ii) other disorders of the nervous system.

Given the convergence of our neuroimaging and genetic associations on movement related circuits of the brain and movement disorders, p.C282Y homozygosity may lead to brain pathology that is undetected in sub-clinical HH cases. As the treatment of care for disorders like Parkinson’s differs from that of hemochromatosis^3,20^, we looked at the overlap of individuals with neurological diagnoses and a clinical diagnosis of hemochromatosis for p.C282Y homozygote males. We found that for p.C282Y homozygote males with a movement disorder diagnosis (31 individuals), most (24 individuals) did not have a concurrent HH diagnosis. We also found a similar pattern for other nervous system disorders where 10 out of 33 men had a concurrent hemochromatosis diagnosis – see Figure 2 panel B.

## Discussion

We found the most prominent genetic risk factor for HH, p.C282Y homozygosity, was associated with substantially lower T2 intensities in brain regions related to motor control - consistent with iron deposition in these regions. Furthermore, p.C282Y homozygosity in males was associated with increased risk for movement-related disorders and other disorders of the nervous system, but not gait disorders. These results are consistent with both previous case reports of movement disorders in HH individuals^10,11^, as well as a higher disease burden for p.C282Y homozygote males vs. females^2,3^. Moreover, we found that most p.C282Y homozygote males did not have a concurrent hemochromatosis diagnosis when diagnosed with either i) a movement disorder or ii) other disorders of the nervous system. This is of importance given the difference in treatment for HH and movement disorders.

These results are consistent with a class of disorders termed ‘Neurodegeneration with Brain Iron Accumulation’ (NBIA) in which rare genetic mutations lead to iron deposition in the basal ganglia^21^. This iron deposition is believed to lead to oxidative damage of these brain regions impairing their function and resulting in movement deficits. Previous conflicting results regarding neurological manifestations of p.C282Y homozygosity^5,6,7,8,9^ have meant that hemochromatosis has traditionally not been included as a cause of NBIA. These conflicting results are probably due to small sample sizes, no stratification based on sex, and biased subject ascertainment. We believe that our study addresses these issues by conducting disease associations in a sample 500 times larger than the previously listed studies, performing sex-stratified analysis, and selecting individuals on genotype, not disease status. Furthermore, our neuroimaging results provides strong support that p.C282Y homozygosity imparts a large, selective effect on the brain’s motor circuits. The results presented here suggest revisiting p.C282Y homozygosity as a form of NBIA, albeit with reduced penetrance.

The globus pallidus is a region that appears to show large amounts of iron deposition in other NBIA disorders^21,22^, with lower T2 intensities except in juvenile forms (‘eye of the tiger’). Additionally, the previous study investigating the p.C282Y homozygosity effect on T2^*^ intensities in the same sample as our study found a small to moderate effect in the pallidum^14^. However, our analysis did not reveal differences in T2 intensities in the globus pallidus(Figure 1). The lack of association in our analysis could be due to i) a genuine lack of iron deposition in the pallidum when compared other NBIA disorders or ii) a result of biological processes having opposing effects on T2 intensity (e.g. iron deposition decreasing intensities and gliosis increasing intensities) averaging out. Differing sensitivities of T2-weighted vs T2^*^-weighted scans to minerals^23^ likely explain the divergence in results of the pallidum between the previous study^14^ and ours. Further research should be conducted to understand processes occurring in the pallidum of p.C282Y homozygotes and if these truly do differ from other NBIA disorders.

Post mortem samples and in-vivo imaging of individuals diagnosed with Parkinson’s display iron deposition in the brain regions we identified^24^. The most recent genome-wide association study (GWAS) of Parkinson’s disease in males did not identify the variant at position p.C282Y as a risk factor (p=0.16)^25^. We hypothesize this is likely due to the study employing a GWAS standard additive model of inheritance, in which an additional copy of a risk allele imparts a dose-dependent risk. Consistent with findings from previous literature of HH^2^, here we tested a recessive model of inheritance and observed a relatively sizeable increased risk for movement-related disorders in general (OR=1.82) and Parkinson’s disease in particular (OR=1.78) – this is in the 5×10^−6^ percentile of effect sizes from the most significant previous GWAS of Parkinson’s disease in males^26^. Further work needs to be done to verify the size and confidence of this effect in other ancestry groups and populations, particularly given the non-uniform distribution of p.C282Y across the globe (see supplementary Figure 3).

Additionally, although we observe largest T2 intensity reductions (compatible with iron accumulation) in male p.C282Y homozygotes, we still observe a reduction in females and a non-significant increase risk for movement disorders. If, as others have argued^3^, menstruation expels iron from the body, we may be observing the post-menopausal accumulation of iron that has not had enough time to impart the toxic effects compared to males. Indeed, the average age of the women in our neuroimaging study was 64 years old. Furthermore, estrogen may be playing an antioxidant role that is moderating the damaging effects of iron overload^27^.

Although our neuroimaging results are consistent with iron accumulation in associated regions of the brain, lower T2 intensities may also indicate calcification. Indeed a recent case study reported an HH individual who displayed both iron and calcium accumulation for the same brain regions discovered in our analysis^28^. One possible explanation for this observed calcium deposition is that calcium and iron homeostasis are reciprocally connected within these regions. This hypothesis is supported by work demonstrating that iron, as Fe2+ ions, can lead to build of inorganic pyrophosphate with calcium^29^. Additional brain imaging of HH and p.C282Y homozygote individuals, using different imaging modalities, may further elucidate the mineral composition of the abnormalities we have observed.

The convergent evidence we have shown consistent with iron deposition in motor circuits of the brain and enrichment for movement and neurological disorders, in those who are homozygous for p.C282Y suggests considering p.C282Y homozygosity as a risk factor for movement disorders in males. Furthermore, given that early treatment of HH has been shown to be effective in preventing the negative health manifestations of the disease outside of the nervous system^3^, our findings provide additional support for revisiting the public health implications of early screening of populations in which this variant is particularly prevalent.

## Data Availability

Data were obtained from the UK Biobank under accession number 27412. Data is available to approved researchers at https://www.ukbiobank.ac.uk.

