## Supplementary Figures and Tables for "Genetic Risk for Hemochromatosis is Associated with Movement Disorders"

### Supplementary Materials

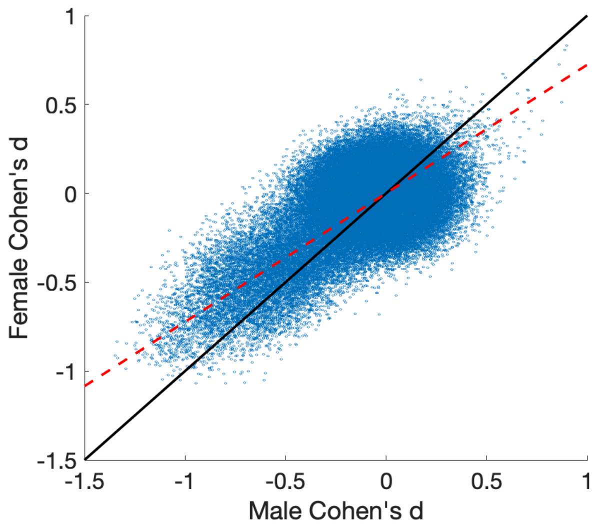


*Figure 1 Sex stratified associations of p.C282Y homozygosity and T2 voxel intensities (Cohen's d), as scatter plot – each line represents a single voxel. Black line indicates y=x (i.e. equal effect sizes). Red line indicates best fitting line (for points of at least moderate effects $x^2 + y^2 > 0.2$), $\beta = 0.72$ indicating approximately a 28% reduction in T2 intensity brain associations for females compared to males.*

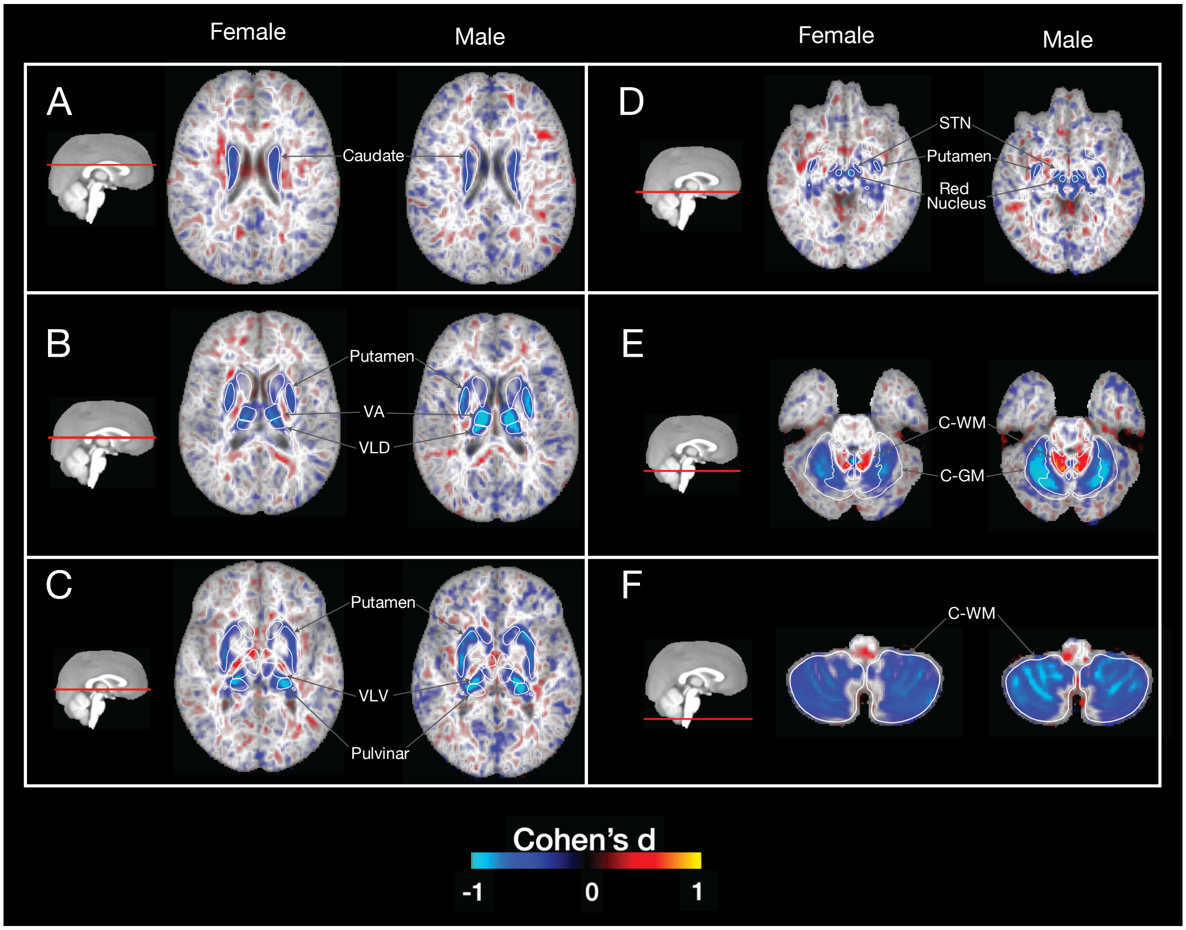


*Figure 2 Maps of sex stratified associations of p.C282Y homozygosity and T2 voxel intensities (Cohen's d). Overall effects are larger in males vs females, with largest effects in males observed in the VA, VLD and pulvinar nuclei of the thalamus (B) as well as in the C-WM and C-GM (E). Abbreviations: VA – ventral anterior, VLD – ventral anterior dorsal, VLV – ventral anterior ventral, C-WM – cerebellum white matter, C-GM – cerebellum GM.*

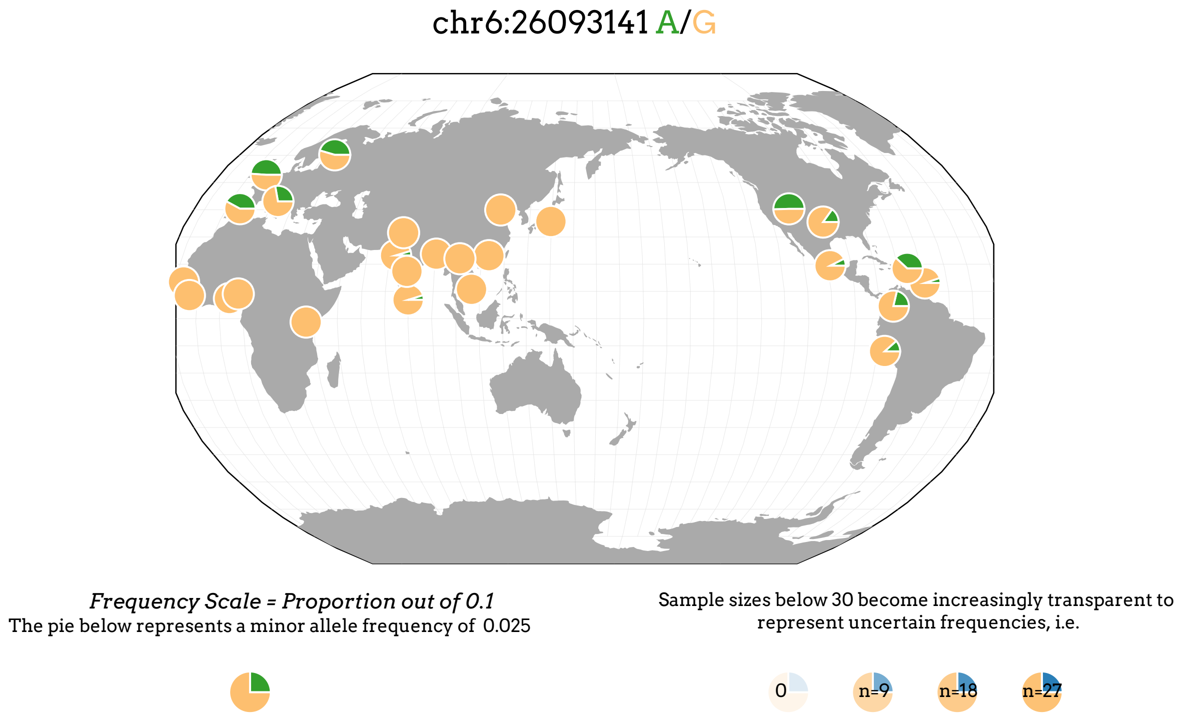


*Figure 3 Global distribution of p.C282Y (rs1800562). Allele frequencies are highest in Europe (particularly northern Europe) and parts of north America. Map generated from the Geography of Genetic Variants Browser[1].*

| FreeSurfer 5.3 segmentation T1 | Pauli, 2018 HCP T1 & T2 | Najdenovska, 2018 HCP FODs |
|---|---|---|
| Amygdala (Amg) | Putamen (Pu) | Anterior (tA) |
| Hippocampus (Hipp) | Caudate (Ca) | Ventral anterior (tVA) |
| Putamen (Pu) | Nucleus accumbens (NAcc) | Mediodorsal (tMD) |
| Caudate (Ca) | Extended amygdala (EA) | Ventral-latero-ventral (tVLV) |
| Globus pallidus (GP) | Substantia nigra pars compacta (SNpc) | Ventral-latero-dorsal (tVLD) |
| Accumbens area (NAcc) | Substantia nigra pars reticulata (SNpr) | Central-latero-lateral-posterior-medial-pulvinar (tC) |
| Thalamus (Thal) | Red nucleus (RN) | Pulvinar (tP) |
| Ventral Diencephalon (VDC) | Parabrachial pigmented nucleus (PBP) | |
| | Hypothalamus (Hyp) | |
| | Mamillary nucleus (MN) | |
| | Subthalamic nucleus (STN) | |

*Table 1* ***Regions of interest (ROIs) labelled using 3 different methods.*** *Column 1) automatic segmentation using FreeSurfer 5.3 applied to each subject's T1 image in atlas space[2]; Column 2) registration of the Pauli atlas of subcortical nuclei to the multispectral atlas[3]; Column 3) registration of the the Najdenovska thalamic nuclei atlas to our data[4].*

*Males*

| Diagnosis | Total Controls | Total Cases | C282Y Cases | OR | CI_lower | CI_upper | P |
|---|---|---|---|---|---|---|---|
| **Abnormalities of Gait and Mobility [R26]** | 219012 | 4557 | 26 | 0.964119 | 0.65175 | 1.4262 | 0.854872 |
| **Other Disorders of Nervous System [G90-99]** | 219737 | 3832 | 33 | 1.5081 | 1.06499 | 2.13558 | 0.02063 |
| **Parkinson's Disease [G20]** | 221638 | 1931 | 20 | 1.78188 | 1.13941 | 2.78661 | 0.011341 |
| **Essential Tremor [G25]** | 222688 | 881 | 10 | 1.92304 | 1.02705 | 3.6007 | 0.0410158 |
| **Movement Disorders [G20-26]** | 220636 | 2933 | 31 | 1.8162 | 1.26605 | 2.60542 | 0.00118983 |
| **Movement and Other Disorders of the Nervous Sy...** | 217030 | 6539 | 61 | 1.63773 | 1.26297 | 2.12371 | 0.00019852 |

*Females*

| Diagnosis | Total Controls | Total Cases | C282Y Cases | OR | CI_lower | CI_upper | P |
|---|---|---|---|---|---|---|---|
| **Abnormalities of Gait and Mobility [R26]** | 260493 | 4226 | 31 | 1.16012 | 0.810591 | 1.66038 | 0.41681 |
| **Other Disorders of Nervous System [G90-99]** | 260857 | 3862 | 25 | 1.06932 | 0.719118 | 1.59005 | 0.740586 |
| **Parkinson's Disease [G20]** | 263551 | 1168 | 6 | 0.79401 | 0.354918 | 1.77633 | 0.574492 |
| **Essential Tremor [G25]** | 263662 | 1057 | 9 | 1.34698 | 0.696958 | 2.60323 | 0.375601 |
| **Movement Disorders [G20-26]** | 262280 | 2439 | 17 | 1.09684 | 0.678124 | 1.77411 | 0.706349 |
| **Movement and Other Disorders of the Nervous Sy...** | 258589 | 6130 | 40 | 1.05775 | 0.771525 | 1.45016 | 0.727282 |

*Table 2 Regression tables results for associating C282Y homozygote (+/+) cases with diagnoses of different neurological disorders. Models were fit separately for males and females.*
